# Active Health Problems in Real-World Consultations on a Purpose-Built Medical AI Platform

**DOI:** 10.64898/2026.11.15.26368158

**Authors:** Blythe Adamson, Aaron W. Li, Sarah Gilson, Bennett J. Waxse, Geetu Ambwani, Byron Crowe, Adam Oskowitz

**Author notes:** **Corresponding author:** Blythe Adamson, PhD, MPH, Doctronic Inc. 1460 Broadway, New York, NY 10036. **Description (submission field, 1-2 sentences):** In 26,000 randomly sampled consultations on a free patient-facing medical AI platform, users presented with concrete clinical problems, led by symptoms such as headache and abdominal pain, and visit intentions were dominated by new or worsening conditions rather than general information seeking, reversing the rank order reported for a general-purpose assistant. More than half of consultations were completed with structured clinical documentation, and 15% of those progressed to a completed physician telehealth visit.

## Abstract

**Importances:** Consumers use conversational artificial intelligence (AI) for health at scale, but whether purpose-built medical AI is used primarily for information seeking or as an entry point into care remains unclear.

**Objective:** To characterize the clinical concepts users present, their visit intentions, and progression to physician telehealth care in real-world consultations on a purpose-built medical AI platform.

**Design:** Exploratory retrospective cohort study of consultations sampled in equal monthly numbers between June 1, 2025, and June 30, 2026.

**Setting:** A free, publicly available, consumer-facing, anonymous-by-default medical AI platform.

**Participants:** Random sample of 26,000 English-language consultations by users aged 18 years or older, each with at least 5 chat interaction rounds.

**Exposure:** A chat consultation with the medical AI.

**Main Outcomes and Measures:** Clinical concepts linked approximately to SNOMED CT; visit intentions classified with a prespecified 11-category schema and reclassified with a published 12-category consumer-health taxonomy; completion of the AI consult with a structured SOAP note; and progression to a completed physician telehealth visit. Automated extraction pipelines were validated against physician annotation in a random subsample.

**Results:** Of 26,000 sampled consultations, 56.7% were by users recorded as female and 50.9% by users aged 18 to 34 years. Among 25,975 consultations with successful extraction, 77,597 clinical concepts were identified; headache, abdominal pain, fatigue, and fever were the most common presenting concepts. Of 55,257 classified intentions, 60.1% (95% CI, 59.7%-60.5%) concerned a new or worsening condition. When consultations were reclassified with the published consumer-health taxonomy, 74.3% (95% CI, 73.8%-74.8%) included symptom questions or health concerns, whereas 5.0% (95% CI, 4.7%-5.2%) included health information or education, reversing the rank order published for a general-purpose assistant. Overall, 53.9% of sampled consultations ended with a structured SOAP note, and 15.1% of those progressed to a completed physician telehealth review.

**Conclusions and Relevance:** Users of this purpose-built medical AI platform predominantly presented active personal health problems rather than general information requests. Symptom-driven use, generation of structured clinical documentation, and progression to physician care suggest that purpose-built medical AI may function as an entry point to clinical care for some users.

**Key Points:** *Question:* Do people use a purpose-built medical AI platform primarily to seek health information or to present active personal health problems?

*Findings:* In this retrospective cohort study of 26,000 randomly sampled consultations on a medical AI platform, 74.3% included symptom questions or health concerns and 5.0% included health information or education, reversing the rank order published for a general-purpose assistant.

*Meaning:* Users predominantly presented active personal health problems rather than general information requests, and purpose-built medical AI may function as an entry point to clinical care for some users.

## Introduction

Consumers have adopted conversational artificial intelligence (AI) for health at scale, as seen in a nationally representative survey conducted in early 2026, 32% of US adults reported using an AI chatbot for health information in the past year.^1^ Adoption is occurring against a backdrop of constrained access to care, with more than 108 million Americans living in designated primary care shortage areas,^2^ and adults without a usual source of primary care are 79% more likely to use chatbots for health information.^3^ Nearly all evidence about this behavior comes from general-purpose AI assistants, and it describes health conversations only in broad categories.^4–7^ Analyses of consumer chat logs,^8^ surveys of adolescents and young adults,^9^ and earlier search-engine query-log research^10^ reach the same conclusion that people bring genuine clinical questions to tools not designed for clinical care.

To our knowledge, no study has described the topics of clinical content in this demand for care. The recent literature has characterized broad domains of health information seeking behaviors, but not the types of symptoms people ask about, the specific conditions they are concerned about, or the relevant clinical specialties. In this study, we characterized 26,000 consultations sampled over 13 months from a patient-facing medical AI platform. We evaluated the specific medical concepts users presented, coded to SNOMED CT, their visit intentions, classified with the same published taxonomy for contextualization with the general-purpose assistant, and rates of progression to a completed physician telehealth visit.

## Methods

### Data source

Doctronic operates a free, publicly available, consumer-facing, anonymous-by-default medical AI platform (www.doctronic.ai) presented as a primary care style consultation. It communicates that the consultation is AI and does not replace a licensed physician. Users complete an AI consult, which is a chat conversation with the medical AI, and receive potential diagnoses, a treatment plan, and a clinically formatted SOAP (Subjective, Objective, Assessment, Plan) note to share with a healthcare provider. Unlike an open-ended general-purpose assistant, the AI consult has a defined endpoint. The AI continues to ask follow-up questions until it has sufficient information to summarize the history, suggest potential diagnoses, and recommend management, and the consult then concludes with generation of the note. A consultation can end without a note if the user enters the diagnostic flow but stops answering its questions before it completes. The AI consult is free at every step. Users may optionally progress from the free AI consult to a telehealth visit with a licensed physician in the affiliated clinic, available in all 50 US states and the District of Columbia, for a fee of$39 that can be paid by the user directly or through health insurance. This exploratory retrospective observational cohort study analyzed de-identified AI consult transcripts and encounter-level operational metadata (Figure 1). This study followed the Strengthening the Reporting of Observational Studies in Epidemiology (STROBE) reporting guideline.

**Figure 1.**
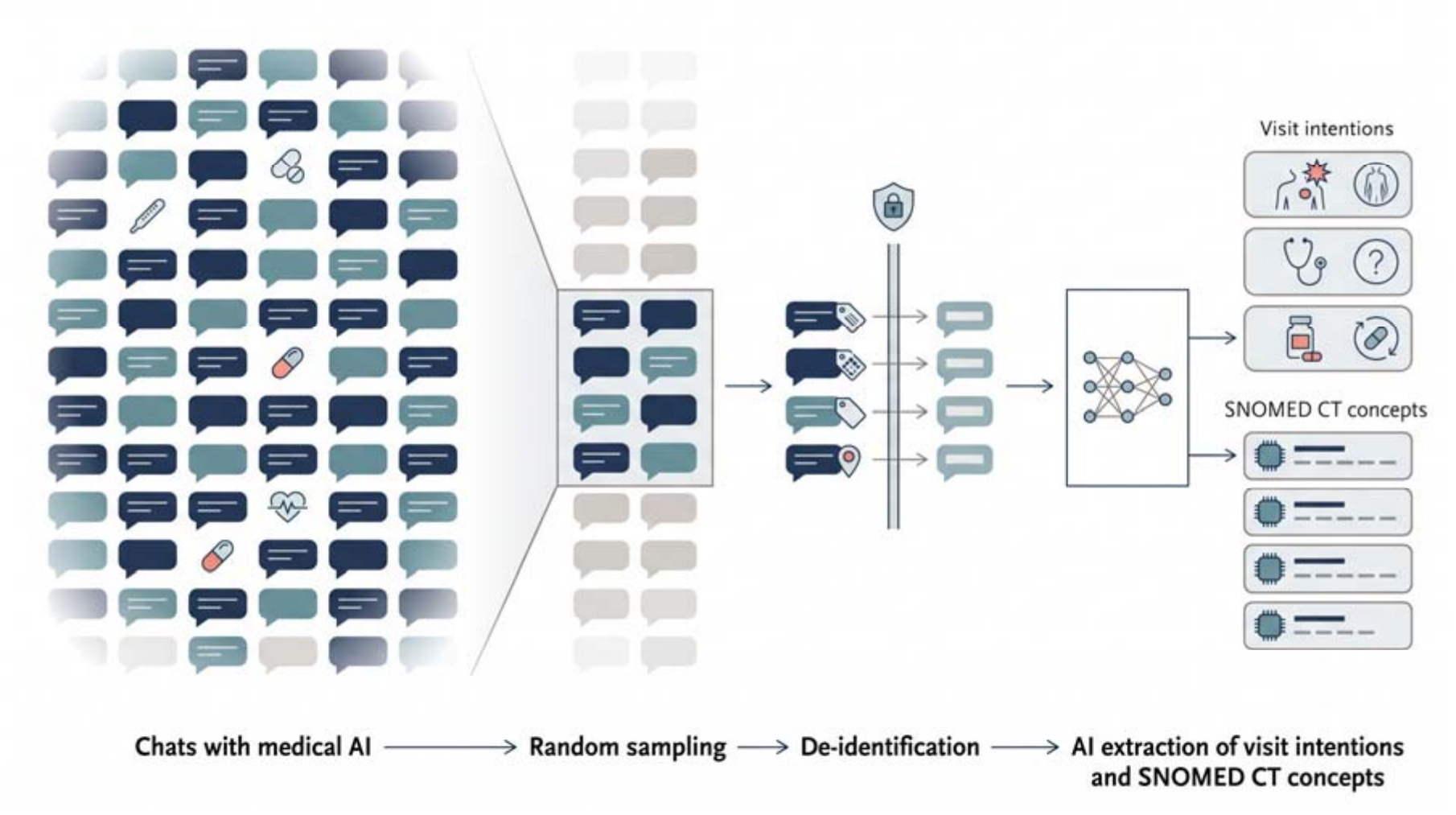
Study Design. Conceptual diagram of the study design: consultations (chats) with the medical AI were randomly sampled, de-identified, and processed by a large language model extraction pipeline to identify visit intentions and SNOMED CT concepts. SNOMED CT indicates Systematized Nomenclature of Medicine, Clinical Terms.

### Population sample

Eligible for sampling were individuals aged 18 years or older who engaged in chat with the medical AI between June 1, 2025, and June 30, 2026 (13 calendar months). We excluded consults with fewer than five chat interaction rounds (26%), a threshold ensuring sufficient conversational content for concept and intention extraction, given briefer exchanges often do not contain enough text to determine intent. From this population we drew a random sample of 2,000 consultations per month (26,000 in total). Sampling managed the computational cost of extraction and represented each month equally because the platform grew rapidly over the study window. The sample was global; device time zone region served as a coarse geographic indicator. Age and sex were user-entered intake information. Consultations conducted in a language other than English were excluded.

### Medical concept extraction

For each sampled consultation, a pipeline extracted clinical concepts from the de-identified conversation text and linked each concept to the Systematized Nomenclature of Medicine, Clinical Terms (SNOMED CT)^11^ by lexical fuzzy matching, returning a concept type, clinical status, and match score against a prespecified threshold; concepts below the threshold were retained in tabulations. Per protocol, the SNOMED CT linkage is approximate supporting detail rather than validated clinical coding.

### Visit intention extraction

A structured-output large language model (LLM) pipeline identified each distinct visit intention expressed in the de-identified conversation text and returned one or more intent categories per intention from a prespecified set of 11 developed by the study authors (SG, AWL). Intent was inferred primarily from the opening rounds of the conversation. The pipeline was built on the GPT-4o mini model (OpenAI); the output schema was constrained to fixed enumerations, with no training or fine-tuning on study data.

To enable a rough comparison with the published characterization of a general-purpose assistant, the same consultations were reclassified using the 12-category consumer-health taxonomy of Costa-Gomes et al, with the published category definitions embedded verbatim in the classification prompt.^4^

### Validation against physician annotation

Three physician authors (SG, BJW, BC) independently annotated 200 randomly sampled study consultations, each reviewed by 1 physician on the scrubbed transcript. Physicians assigned an intent category and a taxonomy category using the pipelines’ category definitions and rated the assigned SNOMED CT concept as clinically reasonable or not appropriate. Physician labels were the reference standard and agreement is reported as exact-match concordance with exact 95% CIs.

### Privacy considerations

All data were de-identified before research use. Free-text transcripts underwent automated scrubbing that detects and removes personally identifiable information, including names, contact details, government-issued identifiers, and financial account details. All extraction pipelines operated on the anonymous and/or de-identified, scrubbed conversation text. In the primary analyses, no human researcher accessed conversation content. For the validation subsample, physician authors reviewed scrubbed, de-identified transcripts under the same access controls.

Encounter-level data elements were limited to coarse, non-identifying attributes, with ages reported in bands. De-identification and automated scrubbing occurred within Doctronic-controlled systems with access controls. No attempt was made to re-identify users, and results are reported only in aggregate. All data used in this research were collected and used in accordance with the terms of agreement with users, including as set out in the Doctronic privacy policy.^12^

### Statistical analysis

All analyses were descriptive and exploratory. Distributions are reported as counts and percentages with 95% Wilson score confidence intervals on the pooled cohort over the full study window. Taxonomy categories were tabulated at the consultation level; a consultation can express more than 1 category, so shares do not sum to 100%. Consultations whose extraction failed schema validation were excluded, with counts reported. An AI-generated SOAP note served as the surrogate endpoint for completion of the AI visit. For consultations with a SOAP note, we report progression to a telehealth visit at which a Doctronic Physician Group physician reviewed the note, ascertained as of July 14, 2026, with no fixed follow-up window, so later consultations had less time to progress, biasing the estimate downward. Only visits with the affiliated clinic are observable. Users may instead take the SOAP note to an outside clinician, so the estimate understates total progression to clinician care. The platform sets no target for this rate. Because each month contributed 2,000 consultations regardless of volume, pooled estimates describe a month-balanced sample rather than the volume-weighted consultation population. Analyses used Python 3.13 (Python Software Foundation) with pandas and NumPy; validation analyses used R.

## Results

Sex, age category, and region of the 26,000 sampled medical AI consultations are reported in Table 1. Among the sampled consultations, 53.9% (95% CI, 53.2%-54.5%) completed the AI consult with a SOAP note summary, and 15.1% (95% CI, 14.6%-15.7%) of those with a SOAP note proceeded to a telehealth visit with a physician. Because the affiliated clinic operates only in the United States, this estimate also includes non-US users who could not progress to the affiliated telehealth clinic.

**Table 1.** Baseline Characteristics of Sampled Medical AI Consultations, June 2025 to June 2026.

| Characteristic | n | Denominator | % | 95% CI |
|---|---|---|---|---|
| Female | 14,740 | 26,000 | 56.7% | 56.1%-57.3% |
| Age category |  |  |  |  |
| 18 to 34 years | 13,247 | 26,000 | 50.9% | 50.3%-51.6% |
| 35 to 49 years | 6,947 | 26,000 | 26.7% | 26.2%-27.3% |
| 50 to 64 years | 3,864 | 26,000 | 14.9% | 14.4%-15.3% |
| 65 years or older | 1,942 | 26,000 | 7.5% | 7.2%-7.8% |
| Time zone region |  |  |  |  |
| Americas | 15,059 | 19,711 | 76.4% | 75.8%-77.0% |
| Asia | 2,758 | 19,711 | 14.0% | 13.5%-14.5% |
| Europe | 1,138 | 19,711 | 5.8% | 5.5%-6.1% |
| Africa | 430 | 19,711 | 2.2% | 2.0%-2.4% |
| Other | 326 | 19,711 | 1.7% | 1.5%-1.8% |
| Missing or unresolvable | 6,289 | 26,000 | 24.2% |  |
| SOAP note generated (visit completion) | 14,002 | 26,000 | 53.9% | 53.2%-54.5% |

Time zone region is tabulated among the 19,711 consultations with a resolvable device time zone; consultations with a missing or unresolvable time zone are excluded from that denominator and shown in the Missing or unresolvable row against the full sample. Sex was user-entered intake information; the 11,260 consultations not recorded as female were recorded as male (0 with missing sex).

### Medical concepts presented in consultations

The SNOMED CT pipeline extracted 77,597 clinical concepts from 25,975 consultations (25 extraction failures excluded), a mean of 3.0 concepts per consultation. The largest clinical family topic areas were gastrointestinal (15.4% of consultations), musculoskeletal and limb (15.0%), neurologic (13.2%), general and constitutional (13.0%), and sexual and reproductive health (12.6%), with skin, hair, and nail concerns close behind (10.4%). Presenting symptoms led the demand from headache, abdominal pain, fatigue, and fever as the most frequently extracted concepts. Some clinical topic areas were distributed with a long tail of diverse clinical concepts (eg, sexual and reproductive health), while other areas focused on a few specific concepts (eg, neurologic) (Figure 2). The mean lexical match score was 0.95, with 2.8% of concepts below the prespecified 0.7 threshold.

**Figure 2.**
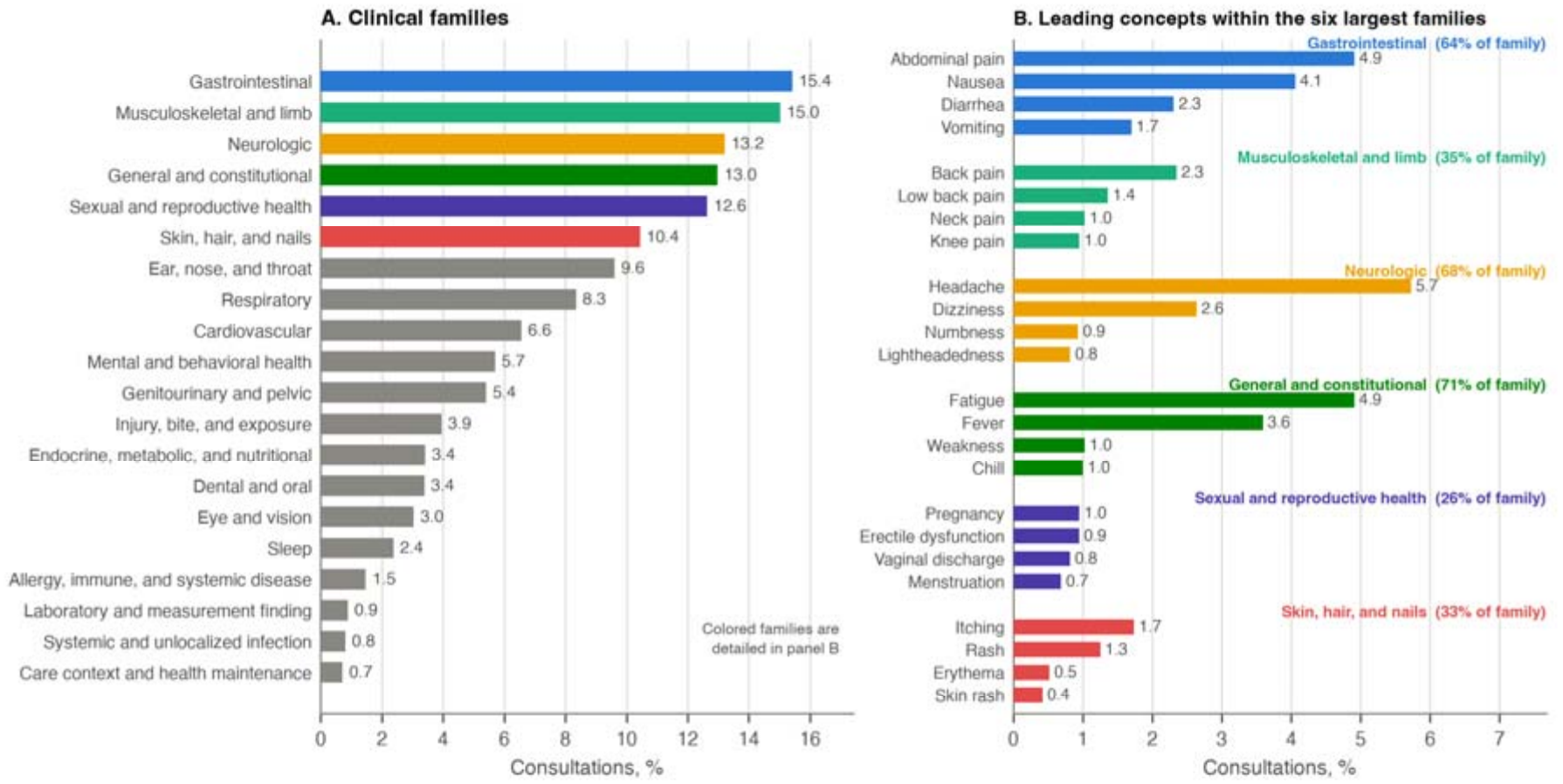
Most Frequent Medical Concepts Extracted From Medical AI Consultations, Coded to SNOMED CT. Panel A shows clinical families as the share of 25,975 processed consultations with at least 1 concept in the family; a consultation can contribute to more than 1 family, so percentages do not sum to 100. Two residual buckets are omitted from panel A: nonspecific or unlocalized findings (11.0%) and the long tail of individually rare concepts (16.3%). Panel B shows the leading concepts within the 6 largest families, as the share of consultations. SNOMED CT indicates Systematized Nomenclature of Medicine, Clinical Terms.

### Visit intentions

The intent extraction pipeline processed 25,975 consultations (25 extraction failures excluded) and identified 55,348 distinct visit intentions (median, 2 per consultation [IQR, 1-3]); 91 unclassifiable intentions were excluded from denominators. A new or worsening condition was the most common first-listed intention, accounting for 60.1% (95% CI, 59.7%-60.5%) of 55,257 classified intentions, followed by health education requests (17.8% (95% CI, 17.5%-18.1%)) and reassurance seeking (7.7% (95% CI, 7.5%-7.9%)); the distribution across all 11 prespecified categories is shown in Figure 3.

**Figure 3.**
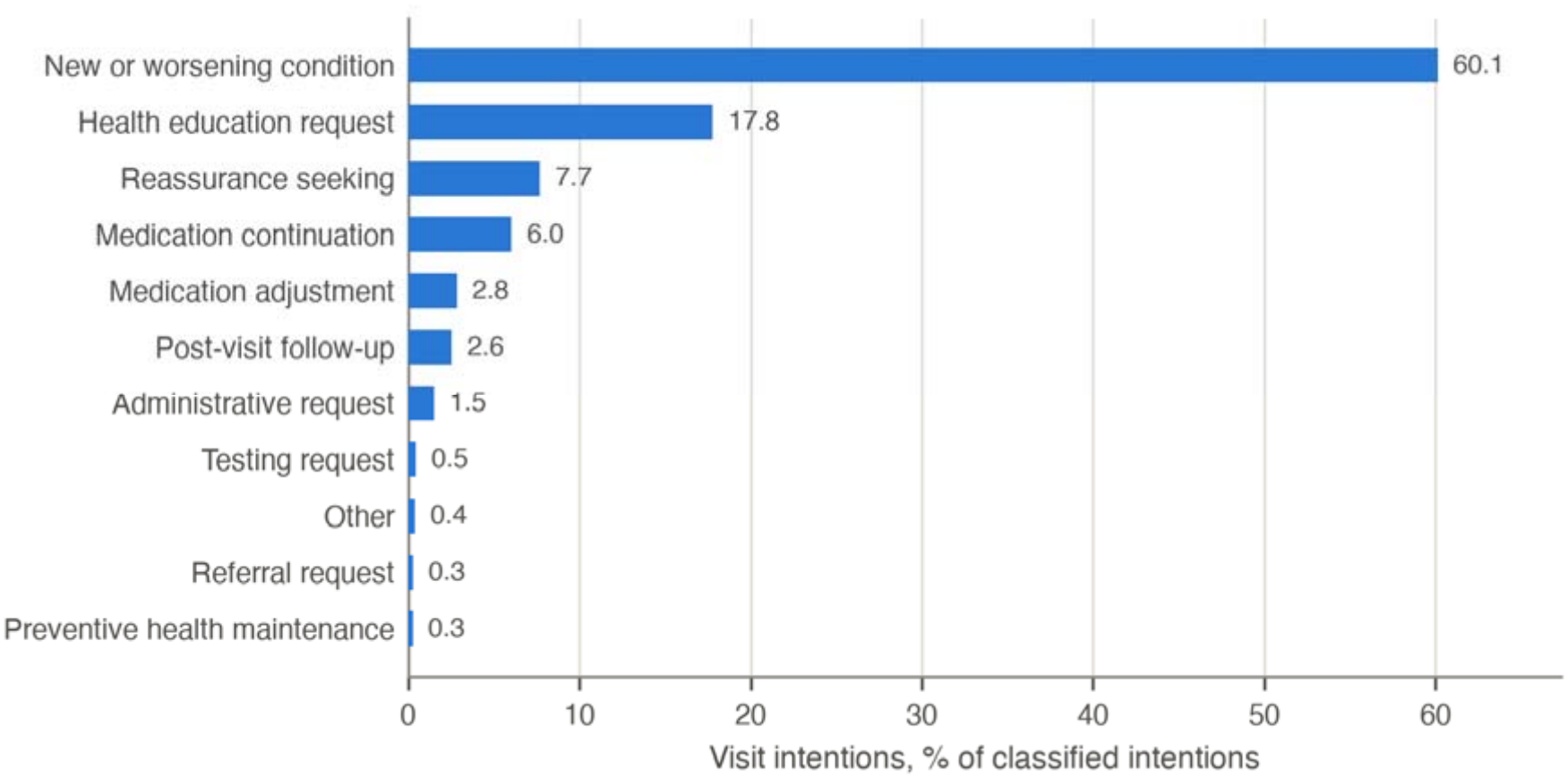
Visit Intentions Extracted From Medical AI Consultations. First-listed intent category per intention, under the study’s prespecified 11-category taxonomy developed by the study authors (SG, AWL), as a percentage of 55,257 classified intentions.

When the same consultations were reclassified with the published 12-category consumer-health taxonomy, the rank order of categories reversed relative to that published for the general-purpose assistant. We found 74.3% (95% CI, 73.8%-74.8%) of 25,998 classified consultations (2 classification failures excluded) included at least 1 intention in Symptom Questions and Health Concerns, whereas Health Information and Education, the leading category on the general-purpose assistant (40.7% of Copilot health-related conversations), appeared in only 5.0% (95% CI, 4.7%-5.2%). Categories designed to capture professional and administrative uses of a general-purpose assistant (Research and Academic Support, Medical Paperwork, Digital Tools and Fitness Apps) were sparsely populated. The full contextualization is presented in Table 2.

**Table 2.** Consultation-Level Intent Distribution for a Patient-Facing Medical AI Platform and a General-Purpose Assistant.

| Taxonomy category | Medical AI platform, n | Medical AI platform, % (95% CI) | General-purpose assistant, % |
|---|---|---|---|
| Symptom Questions and Health Concerns | 19,320 | 74.3% (73.8%-74.8%) | 10.9 |
| Condition Information and Care Questions | 5,515 | 21.2% (20.7%-21.7%) | 8.0 |
| Health Information and Education | 1,290 | 5.0% (4.7%-5.2%) | 40.7 |
| Other Health and Fitness Intent | 949 | 3.7% (3.4%-3.9%) | NR |
| Medical Paperwork | 830 | 3.2% (3.0%-3.4%) | 9.8 |
| Emotional Well-Being | 810 | 3.1% (2.9%-3.3%) | 3.9 |
| Healthcare Navigation and Access to Care | 632 | 2.4% (2.3%-2.6%) | 3.1 |
| Fitness, Lifestyle, and Coaching | 388 | 1.5% (1.4%-1.6%) | 9.0 |
| Not Health | 203 | 0.8% (0.7%-0.9%) | NR |
| Coverage and Benefits | 157 | 0.6% (0.5%-0.7%) | 2.7 |
| Research and Academic Support | 20 | 0.1% (0.0%-0.1%) | 11.6 |
| Digital Tools and Fitness Apps | 17 | 0.1% (0.0%-0.1%) | 0.4 |

Taxonomy categories and definitions are from Costa-Gomes et al.<sup>4</sup> Medical AI platform values are the share of 25,998 classified Doctronic consultations expressing at least 1 intention in the category; because consultations can express intentions in more than 1 category, shares do not sum to 100%. General-purpose assistant values are the published shares of Microsoft Copilot health-related conversations reported by Costa-Gomes et al and were not recomputed. NR indicates the category was not reported in the published figure. The published values distribute conversations across categories summing to approximately 100%, whereas the medical AI platform values are multi-label prevalences; the two columns therefore support comparison of rank order rather than magnitude. Multi-label tabulation can only increase a category’s share, so the small share for Health Information and Education is conservative with respect to this difference.

### Validation against physician annotation

Among the 200 annotated consultations, pipeline-physician concordance was 73.5% (95% CI, 66.8%-79.5%) for the 11-category intent classification and 79.0% (95% CI, 72.7%-84.4%) for the 12-category taxonomy, comparable to the source study’s 84%, with disagreements concentrated in adjacent categories. Physicians rated the assigned SNOMED CT concept clinically reasonable in 94.8% (95% CI, 90.4%-97.6%) of consultations with an assigned concept.

## Discussion

Users of a purpose-built, consumer-facing medical AI platform presented with concrete clinical problems and care-seeking intent. The most frequently extracted concepts were presenting symptoms such as headache and abdominal pain, and 3 of every 5 first-listed intentions concerned a new or worsening condition. When re-classified with the same published taxonomy used to characterize a general-purpose assistant, we found that information seeking - dominant on the general-purpose assistant - was a small minority on the medical AI platform, where symptom questions and active health concerns dominated. The encounters also connected to downstream clinical care: more than half of sampled consultations generated structured clinical documentation, and 15.1% of those proceeded to a completed physician review during a telehealth visit, an observable lower bound on progression to clinician care. The transferable lesson is that the platform a person chooses may reflect their intent. People may bring general health questions to general-purpose assistants and active personal health problems to medical AI. Interactions on purpose-built platforms may function more like traditional care encounters and may warrant the evidence standards applied to other care settings.

This study has limitations. First, the SNOMED CT linkage is lexical and approximate. Physicians judged the assigned concepts clinically reasonable in most annotated consultations, but the linkage remains supporting detail rather than validated coding. Second, the population of self-selected users of a free, anonymous platform may not be representative of the general population, and minimal demographics are collected by anonymous-by-default design, so representativeness cannot be fully assessed in this cohort. Relatedly, the eligibility threshold of five chat interaction rounds excludes brief exchanges, which are the most likely to be purely informational. The platform returns potential diagnoses and management recommendations only when the consult completes, users seeking a quick informational answer are likely to stop early, so the small share of information seeking partly reflects the eligibility criterion as well as the platform. In a sensitivity analysis, 2,000 excluded consultations with fewer than five interaction rounds were processed with the same extraction pipelines. Relative to the analytic cohort, health education requests were more common (46% vs 30%, with months weighted equally) and new or worsening conditions less common (39% vs 68%), although symptom questions and health concerns remained the most common taxonomy category (51% vs 74%). Third, the contextualization with the prior study is a population-level contrast across platform, period, and classification pipeline rather than a measured effect; the published estimates could not be recomputed because the source data are not available. Automated classification agreed substantially with physician annotation, but residual misclassification is possible, so the contrast supports rank order rather than magnitude. Fourth, the anonymous-by-default design prevents linking consultations from the same individual, so a person may contribute more than 1 consultation and confidence intervals, which assume independence, may be narrow. Finally, this analysis characterizes the demand users brought to the platform, not the clinical quality of the medical AI’s responses. Future studies from this platform will evaluate whether escalation and referral occur when clinically indicated.

In these consultations, care seeking was dominated by active personal health problems that produced structured clinical documentation and linked to downstream physician care. These findings suggest that purpose-built medical AI may function as an entry point to care for some users.

## Supporting information

STROBE Checklist

## Data Availability

Aggregate-level data analyzed in the present study are available upon reasonable request to the authors

## Declarations

### Access to data

Dr Adamson had full access to all the data in the study and takes responsibility for the integrity of the data and the accuracy of the data analysis.

### Conflicts of interest

Authors (BA, AL, SG, BW, GA, BC, AZO) hold equity in Doctronic

Inc. Author BC participated in this study in his capacity as a Doctronic employee and outside of his duties and responsibilities at Stanford.

### Code availability

The analysis code is not publicly available. It was developed within Doctronic’s internal infrastructure and relies on proprietary data pipelines and access-controlled systems that cannot be reproduced externally.

### Data sharing statement

De-identified individual consultation data, including conversation transcripts, cannot be shared because of the privacy protections and terms of agreement with users under which they were collected. Aggregate data supporting the findings are available in the article and its tables.

## Acknowledgment

Generative AI was used in manuscript preparation: Claude Fable 5 (Anthropic, San Francisco, CA) was used for editing the text of this manuscript between July and October 2026, under the direction of the authors. Gemini Nano Banana (Google) was used by BA for generation of the Figure 1 conceptual diagram. The authors reviewed all AI-edited text and take full responsibility for the integrity and accuracy of the content.

