## Supplementary material for "Active Health Problems in Real-World Consultations on a Purpose-Built Medical AI Platform": STROBE Checklist

### STROBE Statement: Checklist of items for cohort studies

**Manuscript:** Active Health Problems in Real-World Consultations on a Purpose-Built Medical AI Platform (DOC-RWE-2026-001, manuscript v0.8, 2026-10-29)

| Item No | Recommendation | Location in manuscript |
|---|---|---|
| **Title and abstract** | | |
| 1a | Indicate the study's design with a commonly used term in the title or the abstract | Abstract, Design ("Exploratory retrospective cohort study") |
| 1b | Provide in the abstract an informative and balanced summary of what was done and what was found | Abstract (structured; Importance through Conclusions and Relevance) |
| **Introduction** | | |
| 2 | Explain the scientific background and rationale for the investigation being reported | Introduction, paragraphs 1-2 |
| 3 | State specific objectives, including any prespecified hypotheses | Introduction, final paragraph (descriptive objectives; no hypotheses, study is exploratory) |
| **Methods** | | |
| 4 | Present key elements of study design early in the paper | Methods, Data source (final sentences) and Abstract, Design |
| 5 | Describe the setting, locations, and relevant dates, including periods of recruitment, exposure, follow-up, and data collection | Methods, Data source (platform description) and Population sample (June 1, 2025 to June 30, 2026); Statistical analysis (progression ascertained as of July 14, 2026) |
| 6a | Give the eligibility criteria, and the sources and methods of selection of participants; describe methods of follow-up | Methods, Population sample (age 18+, English language, at least 5 chat rounds, equal monthly random sampling); Statistical analysis (progression ascertainment) |
| 6b | For matched studies, give matching criteria and number of exposed and unexposed | Not applicable (no matching) |

| Item No | Recommendation | Location in manuscript |
|---|---|---|
| 7 | Clearly define all outcomes, exposures, predictors, potential confounders, and effect modifiers; give diagnostic criteria, if applicable | Methods: Medical concept extraction, Visit intention extraction, Statistical analysis (SOAP note as surrogate completion endpoint; progression definition) |
| 8 | For each variable of interest, give sources of data and details of methods of assessment (measurement); describe comparability of assessment methods if there is more than one group | Methods: Data source (operational metadata, user-entered intake), extraction subsections (pipelines, models), Validation against physician annotation |
| 9 | Describe any efforts to address potential sources of bias | Methods, Statistical analysis (no fixed follow-up window, downward bias stated; month-balanced sampling); Discussion, limitations (eligibility threshold, misclassification; short-chat sensitivity analysis) |
| 10 | Explain how the study size was arrived at | Methods, Population sample (2,000 consultations per month, 26,000 total; rationale: computational cost of extraction and equal monthly representation) |
| 11 | Explain how quantitative variables were handled in the analyses; describe which groupings were chosen and why | Methods, Population sample (ages in bands) and Statistical analysis (counts, percentages, Wilson 95% CIs) |
| 12a | Describe all statistical methods, including those used to control for confounding | Methods, Statistical analysis (descriptive and exploratory only; no confounding adjustment, none claimed) |
| 12b | Describe any methods used to examine subgroups and interactions | Not applicable (no subgroup or interaction analyses) |
| 12c | Explain how missing data were addressed | Methods, Statistical analysis (schema-validation failures excluded with counts reported); Results (failure counts); Table 1 (missing time zone and sex rows) |

| Item No | Recommendation | Location in manuscript |
| --- | --- | --- |
| 12d | If applicable, explain how loss to follow-up was addressed | Methods, Statistical analysis (ascertainment date with no fixed follow-up window; downward bias stated) |
| 12e | Describe any sensitivity analyses | Discussion, limitations (short-chat sensitivity analysis of 2,000 excluded consultations with fewer than 5 rounds) |
| **Results** | | |
| 13a | Report numbers of individuals at each stage of study | Results, opening paragraph and extraction subsections (sampled, processed, extraction failures, classified intentions) |
| 13b | Give reasons for non-participation at each stage | Methods, Population sample (exclusions); Results (extraction and classification failure counts) |
| 13c | Consider use of a flow diagram | No flow diagram; stage counts are reported in text (Figure 1 is a conceptual design diagram) |
| 14a | Give characteristics of study participants and information on exposures and potential confounders | Table 1 (sex, age bands, time zone region, SOAP completion) |
| 14b | Indicate number of participants with missing data for each variable of interest | Table 1 (missing or unresolvable time zone row; missing sex count in the table footnote) |
| 14c | Summarise follow-up time | Methods, Statistical analysis (ascertained as of July 14, 2026; no fixed window) |
| 15 | Report numbers of outcome events or summary measures over time | Results (SOAP completion, progression to telehealth visit, concept counts, intention counts) |
| 16a | Give unadjusted estimates and, if applicable, confounder-adjusted estimates and their precision | Results and Tables 1-2 (unadjusted descriptive estimates with 95% Wilson CIs; no adjusted analyses) |
| 16b | Report category boundaries when continuous variables were categorized | Table 1 (age bands) |

| Item No | Recommendation | Location in manuscript |
|---|---|---|
| 16c | If relevant, consider translating estimates of relative risk into absolute risk | Not applicable (no relative risk estimates) |
| 17 | Report other analyses done | Results, Validation against physician annotation; Results, taxonomy reclassification (Table 2); Discussion, short-chat sensitivity analysis |
| **Discussion** | | |
| 18 | Summarise key results with reference to study objectives | Discussion, paragraph 1 |
| 19 | Discuss limitations of the study, taking into account sources of potential bias or imprecision | Discussion, limitations paragraph (lexical SNOMED linkage, self-selection, eligibility threshold, cross-study contrast, independence assumption) |
| 20 | Give a cautious overall interpretation of results | Discussion, paragraphs 1 and closing ("may function as an entry point to clinical care for some users") |
| 21 | Discuss the generalisability (external validity) of the study results | Discussion, limitations (self-selected users of a free anonymous platform; representativeness) |
| **Other information** | | |
| 22 | Give the source of funding and the role of the funders | Declarations, Funding and Conflicts of interest |
